# Comparison of risk factors for coronary heart disease morbidity versus mortality

**DOI:** 10.1101/19004994

**Authors:** G. David Batty, Mika Kivimäki, Steven Bell

**Affiliations:** Department of Epidemiology and Public Health, University College London, London, UK; School of Biological and Population Health Sciences, Oregon State University, Corvallis, USA; Department of Public Health and Primary Care, University of Cambridge, Cambridge, UK

## Abstract

Owing to the often prohibitively high costs of medical examinations, or an absence of infrastructure for linkage of study members to morbidity registries, much aetiological research in the field of cardiovascular research relies on death records. Because they are regarded as being more distal to risk factor assessment than morbidity endpoints, mortality data are generally maligned in this context for seemingly providing less clear insights into aetiology. The relative utility of mortality versus morbidity registries is, however, untested. In a pooling of data from three large cohort studies whose participants had been linked to both death *and* morbidity registries for coronary heart disease, we related a range of established and emerging risk factors to these two methods of ascertainment. A mean duration of study member surveillance of 10.1 years (mortality) and 9.9 years (morbidity) for a maximum of 20,956 study members (11,868 women) in the analytical sample yielded 289 deaths from coronary heart disease and 770 hospitalisations for this condition. The direction of the age- and sex-adjusted association was the same for 21 of the 24 risk factor– morbidity/mortality combinations. The only marked discordance in effect estimates, such that different conclusions about the association could be drawn, was for social support, total cholesterol, and fruit/vegetable consumption whereby null effects were evident for selected outcomes. In conclusion, variation in disease definition typically did not have an impact on the direction of the association of an array of risk factors for coronary heart disease.

## Introduction

Despite declining rates, coronary heart disease remains a burdensome cause of death and disability worldwide.^1^ In on-going efforts to identify new environmental and genetic risk factors for coronary heart disease, events based on disease incidence are regarded as being preferable to events based on deaths. Incidence data, which may be derived from record linkage or medical examination in population-based cohort studies, are privileged because of their proximity to risk factor assessment, seemingly providing clearer insights into aetiology. By contrast, mortality data comprise not only the morbid event itself but, in the high probability of survival following a heart attack, prognosis. Owing to the often prohibitively high costs of medical examinations, or an absence of infrastructure for linkage of study members to morbidity registries, most investigators have to rely on death records.^2 3^ In a pooling of data from three large cohort studies whose participants had been linked to death *and* hospital registries for morbidity, for the first time, we assessed the relative utility of each ascertainment method by relating them to a range of established and emerging risk factors.^4^

## Methods

We pooled data from the Scottish Health Surveys which comprise three identical prospective cohort studies, baseline data collection for which took place in 1995, 1998 and 2003. Described in detail elsewhere,^5^ risk factor data were collected using standard, identical protocols. Individuals without a history of heart disease hospitalisation were flagged for mortality using the procedures of the UK NHS Central Registry and hospitalisations using Scottish Morbidity Records database.^3^ Ethical approval for data collection was granted by the London Research Ethics Council, or local research ethics councils. Participants gave informed consent.

## Results

A mean duration of study member surveillance of 10.1 years (mortality) and 9.9 years (morbidity) for a maximum of 20,956 study members (11,868 women) in the analytical sample yielded 289 deaths from coronary heart disease and 770 hospitalisations for this condition. Findings for risk factors known to be causally linked to coronary heart disease are presented in figure 1; results for emerging risk factors and those thought to be non-causally associated are depicted in figure 2.

**Figure 1.**
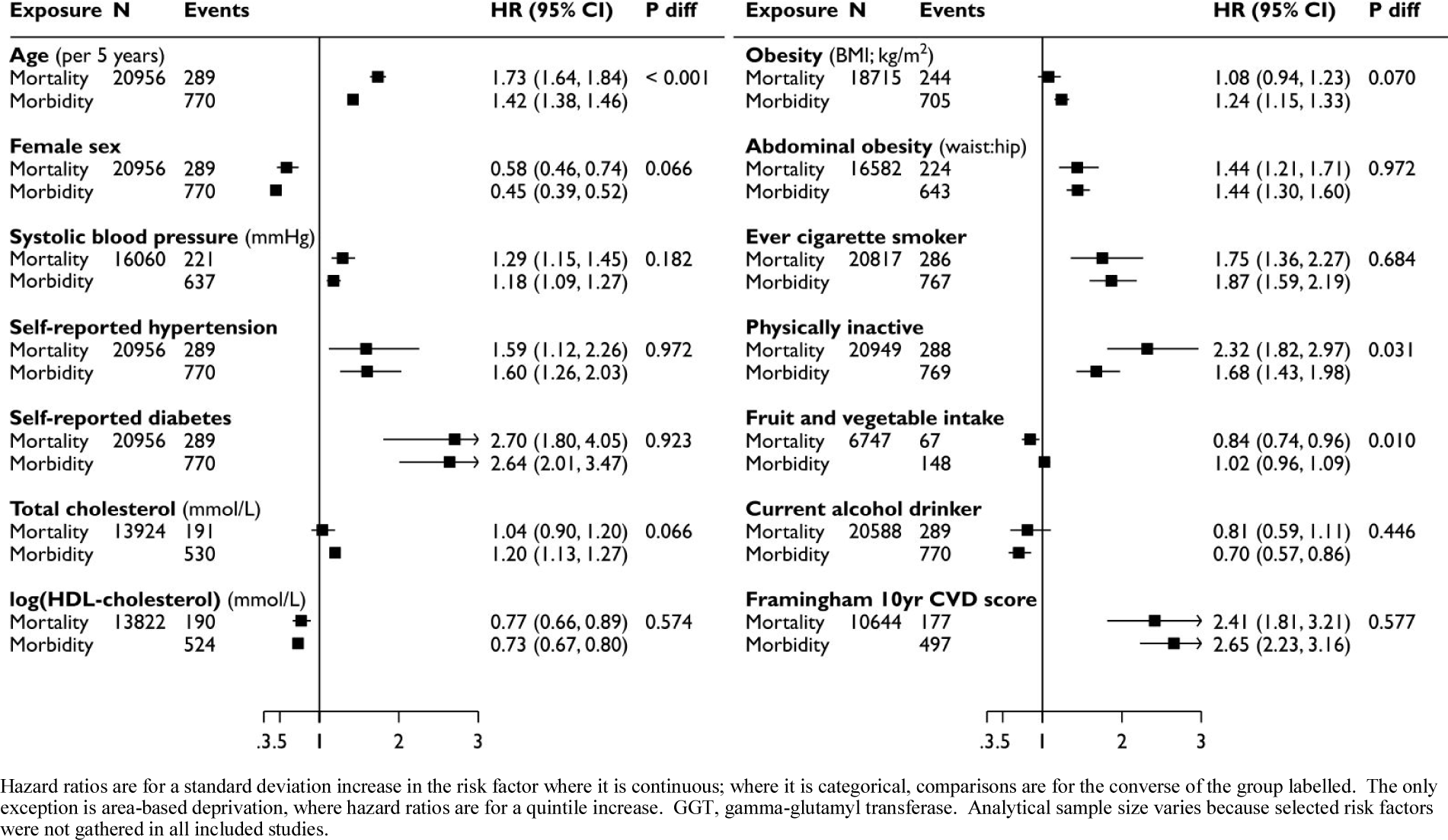
Age- and sex-adjusted hazard ratios (95% confidence interval) for risk factors causally linked to coronary heart disease. Hazard ratios are for a standard deviation increase in the risk factor where it is continuous; where it is categorical, comparisons are for the converse of the group labelled. The only exception is area-based deprivation, where hazard ratios are for a quintile increase. GGT, gamma-glutamyl transferase. Analytical sample size varies because selected risk factors were not gathered in all included studies.

**Figure 2.**
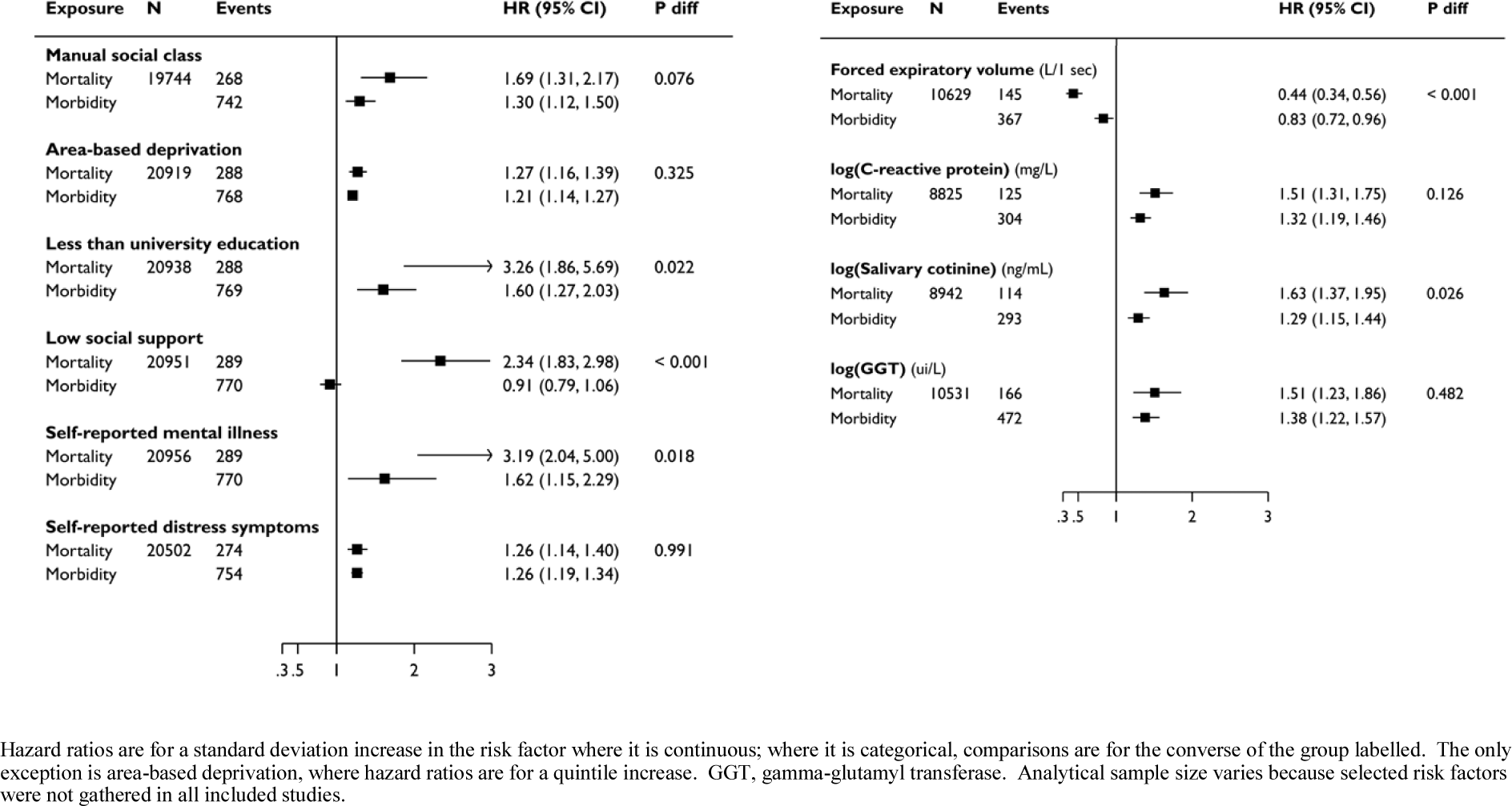
Age- and sex-adjusted hazard ratios (95% confidence interval) for emerging risk factors for, or risk markers that show associations with, coronary heart disease. Hazard ratios are for a standard deviation increase in the risk factor where it is continuous; where it is categorical, comparisons are for the converse of the group labelled. The only exception is area-based deprivation, where hazard ratios are for a quintile increase. GGT, gamma-glutamyl transferase. Analytical sample size varies because selected risk factors were not gathered in all included studies.

The direction of the age- and sex-adjusted association was the same for 22 of the 24 risk factor– morbidity/mortality combinations. As evidenced by the test for heterogeneity by outcome ascertainment, there was, however, occasionally some differences in the magnitude of association, such that somewhat stronger effects were apparent in mortality analyses for age, physical inactivity, (figure 1), educational attainment, mental illness, lung function, and salivary cotinine (a biomarker for cigarette smoke exposure) (figure 2). The only marked discordance in effect estimates, such that different conclusions about the association could be drawn, was for social support (indexed by relationship status), total cholesterol, and fruit and vegetable consumption whereby null effects were evident for selected outcomes. Aggregating risk factors into the Framingham algorithm revealed very similar predictive capacity for coronary heart disease whether based on morbidity or mortality records.

## Discussion

The main finding of the present analyses was that variation in disease definition – morbidity or mortality – did not have an impact on the direction of the association of an array of known risk factors for coronary heart disease. Comparable results reported for another cardiovascular outcome, stroke, support the validity of our findings.^6^

Our findings suggest that for the most common presentation of cardiovascular disease there is no obvious advantage to utilising morbidity records. This has implications for those investigators operating outside countries with well-established data linkage procedures who only have access to death registers. Our findings may also suggest that morbidity data collected via study member attendance at designated clinical research centres have no additional utility, though no such direct comparison was made herein. Lastly, whether morbidity records for other cardiovascular disease sub-types such as peripheral vascular disease, heart failure, abdominal aortic aneurysm, amongst others, also offer no analytical advantage to death records is unknown.

## Data Availability

Data from The Health Surveys for England and The Scottish Health Surveys (https://data-archive.ac.uk/) are available to researchers upon application.

